# Left ventricular strain does not differentiate amyloidogenic profiles in at-risk individuals with *TTR* Val142Ile

**DOI:** 10.1101/2021.01.26.21250565

**Authors:** Amy R. Kontorovich, Wenli Zhao, Michael Gavalas, Helen Hashemi, Steve L. Liao, Eman R. Rashed, Sumeet S. Mitter, Maria Giovanna Trivieri, Stamatios Lerakis, Eimear E. Kenny, Noura S. Abul-Husn

## Abstract

**Objectives:** To identify echocardiographic signatures featuring left ventricular longitudinal strain (LS) associated with genetic risk for cardiac amyloidosis (CA) due to the *TTR* Val142Ile (V142I) variant in African American (AA) and Hispanic/Latinx (H/L) individuals.

**Background:** Hereditary transthyretin amyloidosis (hATTR) can cause CA in ∼60-70% of older V142I carriers, but amyloid deposition progresses over many years. Disease-modifying therapy for CA is now available and early initiation is a priority for improving outcomes. Genomic screening programs and familial cascade genetic testing uncover pre-symptomatic V142I carriers, yet no guidelines exist for early CA detection.

**Methods:** Exome sequencing data linked to electronic health records (EHRs) of Bio*Me* biobank participants were queried for AA or H/L *TTR*- and *TTR*+ (V142I) subjects without hATTR diagnoses and with prior echocardiograms suitable for retrospective LS analysis. Systemic “red flag” features of ATTR were extracted from EHRs of *TTR*+ subjects. Speckle tracking echocardiography was retrospectively applied to determine global (GLS) and segmental LS. Relative apical sparing (RAS) was calculated.

**Results:** 57 *TTR*+ and 46 *TTR*-age- and ancestry-matched subjects were included. GLS declined with age in females but not males, and was abnormal (<16%) in 18 (31.6%) *TTR*+ and 7 (15.2%) *TTR*-subjects (p = 0.066). Apical sparing was observed in 13 (22.8%) *TTR*+ and 11 (23.9%) *TTR*-subjects (p = 1.0). After adjusting for relevant demographic and echocardiographic covariates, neither GLS nor RAS was associated with *TTR*+ V142I status. Red flag features were not associated with GLS or RAS in *TTR*+ subjects.

**Conclusions:** Neither GLS nor RAS were significantly different between *TTR*+ and *TTR*-subjects. Since >50% of *TTR*+ subjects were ≥ 60 years old, penetrance of CA by echocardiography among unselected V142I carriers may be lower than previously estimated. These findings indicate that surveillance for CA in individuals at increased genetic risk due to V142I should not rely solely on echocardiography, even with LS.

---

Hereditary transthyretin amyloidosis (hATTR) due to pathogenic *TTR* variants causes cardiac amyloidosis (CA). Symptoms are frequently insidious and non-specific, resulting in diagnostic delays (1). Since the most common causative variant, Val142Ile (V142I), is prevalent in African American (AA) and Hispanic/Latinx (H/L) individuals (2), missed diagnoses potentiate healthcare disparities. Contemporary diagnostic imaging for CA includes echocardiography, nuclear scintigraphy or SPECT with bone-avid tracers (i.e. technetium-99 pyrophosphate), and cardiac magnetic resonance (1). The utility of these modalities for capturing presymptomatic cardiac changes has not been tested. Echocardiography with left ventricular longitudinal strain (LS) analysis may be well-suited due to its accessibility and low risk. We retrospectively analyzed echocardiograms of *TTR+* and *TTR*-subjects from the multiethnic Bio*Me* Biobank for signatures harkening likely CA onset in those at high genetic risk.

The study was approved by Mount Sinai’s Institutional Review Board (GCO # 19-01925 ISMMS). *TTR*+ subjects were exome-sequenced adults harboring V142I, without diagnostic codes related to hATTR, and with ≥1 echocardiogram in their electronic health record (EHR). *TTR*-subjects were those without pathogenic *TTR* variants, with ≥1 echocardiogram, without diagnostic codes related to hATTR, cardiomyopathy, or heart failure (HF), and matched to *TTR*+ subjects by sex, self-reported race/ethnicity, and age. Post-processing speckle tracking echocardiography (STE, Philips aCMQ) was performed on apical 2-, 3- and 4-chamber views by two blinded independent operators (W.Z., M.G.). Blinded review (S.L.L.) of LS bullseye maps graded presence/absence of apical sparing, and relative apical sparing (RAS) was calculated (3). Echocardiographic parameters were extracted from imaging reports. EHRs were queried for diagnostic codes related to red flag features of hATTR: carpal tunnel syndrome, spinal stenosis, and polyneuropathy (4). Statistical analyses included nonparametric Wilcoxon rank sum or Spearman’s correlation tests.

We identified 57 AA or H/L *TTR*+ subjects without hATTR (including 8 with cardiomyopathy/HF by time of echocardiogram) and 46 well-matched *TTR*-subjects (Figure 1A). Retrospective STE measurements of global LS (GLS) were reproducible (intraclass and interclass correlation coefficients 0.77 and 0.86, respectively). GLS declined with age in females (p = 0.0032) but not males, and was abnormal (<16%) in 18 (31.6%) *TTR*+ and 7 (15.2%) *TTR*-subjects (Figure 1B, p = 0.066). Apical sparing was observed in 13 (22.8%) *TTR*+ and 11 (23.9%) *TTR*-subjects (p = 1.0). Neither GLS (p = 0.37) nor RAS (p = 0.44) was associated with *TTR* status; this held true after adjusting for relevant covariates, and after excluding *TTR*+ individuals with cardiomyopathy/HF. In *TTR*+ subjects, red flag features (present in 27 (47.4%)) were not associated with GLS (p = 0.44) or RAS (p = 0.45).

**Figure 1.**
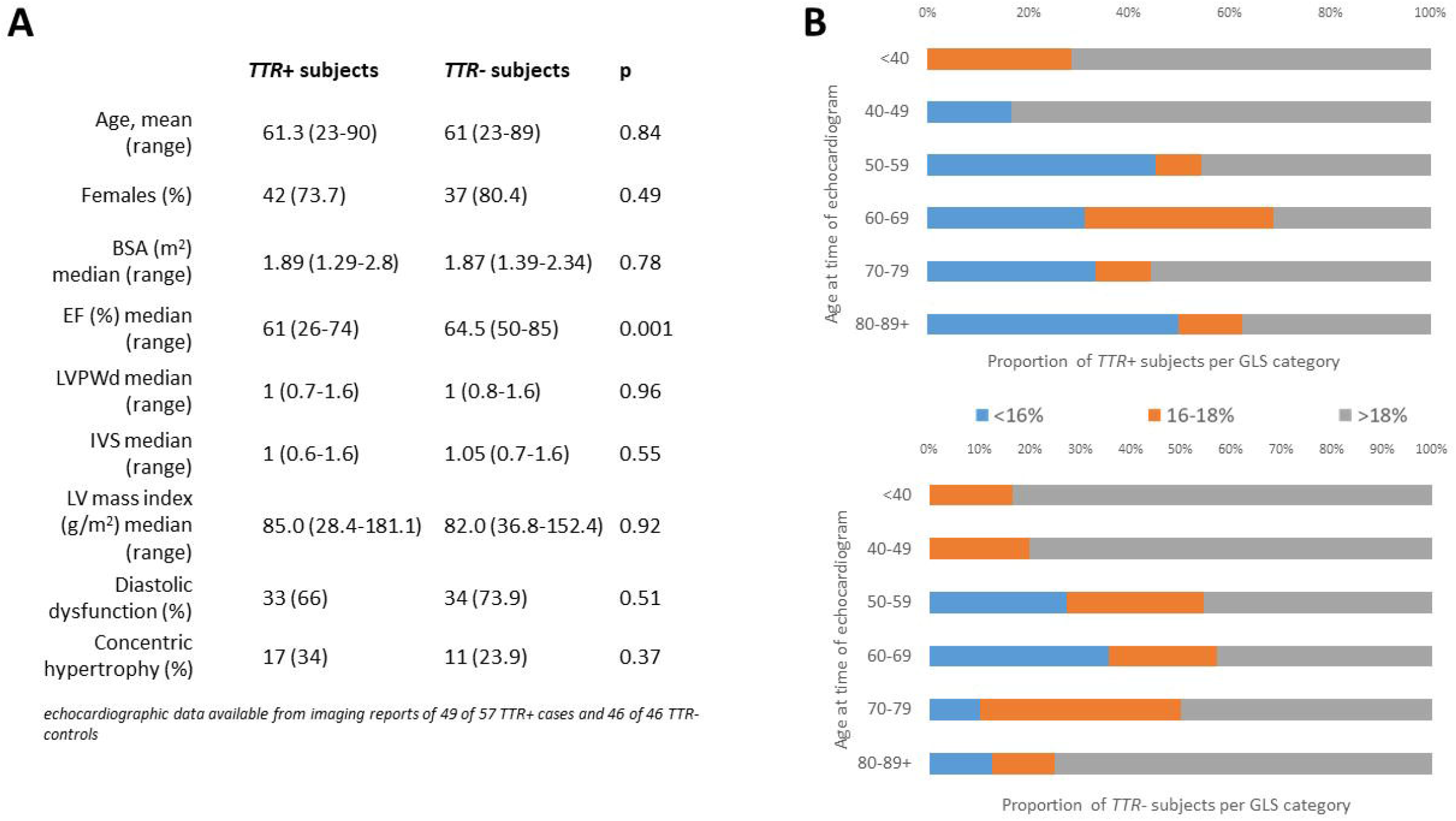
Characteristics of *TTR*+ *and TTR*-subjects. **A)** Subjects were well-matched by demographic and echocardiographic features; **B)** Proportions of *TTR*+ and *TTR*-subjects with abnormal (<16%), borderline (16-18%), and normal (>18%) GLS were similar.

Knowledge of hATTR-related CA is primarily based on traditional phenotype-first studies, which define clinical features in patients with manifest disease. Genotype-first studies of unselected populations afford better understanding of disease penetrance by avoiding phenotype ascertainment bias. The feasibility of detecting preclinical CA in asymptomatic V142I carriers has not been previously evaluated. We found that neither GLS nor RAS distinguished *TTR*+ subjects without hATTR from *TTR*-subjects. CA penetrance in V142I carriers ≥ 65 years is estimated at ∼70-80% (5), yet we observed abnormal strain in only 38.5% of *TTR*+ subjects ≥ 65 years in our cohort, suggesting lower penetrance. Our retrospective study limited the ability to perform longitudinal assessment or evaluate confirmatory endpoints of CA.

Although echocardiography with LS has been proposed as a convenient screening tool for hATTR, it may not identify preclinical CA in individuals at increased genetic risk due to V142I. Still, higher risk assessments (i.e. ionizing radiation from scintigraphy or invasive endomyocardial biopsy) may not be justified, even in individuals with LV wall thickening, abnormal LS, and/or red flags, unless additional clinical features raise suspicion for CA.

## Data Availability

The datasets generated during and/or analyzed during the current study are available from the corresponding author on reasonable request.

